# Factors Associated with Stroke after COVID-19 Vaccination: A Statewide Analysis

**DOI:** 10.1101/2023.02.05.23285486

**Authors:** Fadi Nahab, Rana Bayakly, Mary Elizabeth Sexton, Manet Lemuel-Clarke, Laura Henriquez, Srikant Rangaraju, Moges Ido

## Abstract

**Background:** The objective of our study was to evaluate baseline characteristics, COVID-19 infection and vaccine type and their association with stroke early after COVID-19 vaccination.

**Methods:** In a retrospective cohort study, we estimated the 21-day post vaccination incidence of stroke among COVID-19 first dose vaccine recipients. We linked the Georgia Immunization Registry with the Georgia Coverdell Acute Stroke Registry and the Georgia State Electronic Notifiable Disease Surveillance System data to assess the relative risk of stroke by vaccine type.

**Results:** About 5 million adult Georgians received at least one COVID-19 vaccine from December 1, 2020 to February 28, 2022: 54% received BNT162b2, 41% mRNA-1273 and 5% Ad26.COV2.S. Those with concurrent COVID infection within 21 days post vaccine had an increased risk of ischemic (OR=8.00, 95% CI: 4.18, 15.31) and hemorrhagic stroke (OR=5.23, 95% CI: 1.11, 24.64) with no evidence for interaction between vaccine type and concurrent COVID-19 infection. The 21-day post vaccination incidence of ischemic stroke was 8.14, 11.14, and 10.48 per 100,000 for BNT162b2, mRNA-1273 and Ad26.COV2.S recipients, respectively. After adjusting for age, race, gender, and COVID-19 infection status there was a 57% higher risk (OR=1.57, 95% CI: 1.02, 2.42) for ischemic stroke within 21 days of vaccination associated with the Ad26.COV2.S vaccine compared to BNT162b2.

**Conclusions:** Concurrent COVID-19 infection had the strongest association with early ischemic and hemorrhagic stroke after first dose COVID-19 vaccination. The Ad26.COV2.S vaccine was associated with a higher risk of early post-vaccination ischemic stroke than BNT162b2.

## Introduction

To tackle the emergence of the severe acute respiratory syndrome coronavirus 2 (SARS-CoV-2) virus multiple coronavirus disease 2019 (Covid-19) vaccines have been approved.^1,2^ These vaccines have been shown to be both safe and effective although rare thrombotic events associated with the Ad26.COV2.S vaccine have led to warnings about use in certain high risk populations and a preference for other available vaccines.

The first case of cerebral venous sinus thrombosis (CVST) associated with thrombocytopenia and disseminated intravascular coagulation after Ad26.COV2.S vaccination was documented in 2021.^3–5^ Thrombosis with thrombocytopenia syndrome (TTS) has been reported in 3.83 cases per million doses administered and 0.6 per million deaths and the Advisory Committee on Immunization Practices (ACIP) subsequently performed a comprehensive review of existing data and made a preferential recommendation for mRNA COVID-19 vaccines over the Ad26.COV2.S vaccine.^6^

A systematic review of reported acute ischemic stroke (AIS) early after COVID-19 vaccination identified 31 patients with vaccine-induced immune thrombotic thrombocytopenia (VITT), 95% of whom received viral vector vaccines.^7^ Other studies focused solely on mRNA vaccination have not identified an increased incidence of ischemic stroke in the post-vaccine period.^8,9^ However, large-scale population studies assessing factors associated with stroke risk early after COVID-19 vaccinations are limited outside of case reports and a self-controlled case series method in the French population.^10^ Here we report a statewide analysis to assess baseline factors, COVID-19 infection and vaccination type associated with early incidence of stroke after first dose COVID-19 vaccination.

## Methods

### Study Design and Data Sources

A retrospective cohort study was performed to assess the incidence of acute stroke among COVID-19 vaccine recipients. All adult residents of Georgia who received any type of COVID-19 vaccine from December 1, 2020 to February 28, 2022 were included in the study. The Georgia Registry of Immunization Transactions and Services (GRITS) database served as the source of information on vaccination status, and it includes demographic and personally identifiable information such as date of birth, sex, race and first and last names. The GRITS data was linked with the Georgia Coverdell Acute stroke Registry (GCASR) and the Georgia Sate Electronic Notifiable Disease Reporting System (SENDSS) data of the corresponding time period. The GCASR collects clinical information on patients diagnosed with acute stroke in non-federal hospitals located in Georgia. The GCASR was established in 2005 for monitoring and improving the quality of stroke care across the state and receives data from 83 hospitals which provide service for more than 96% of the stroke patients in Georgia.^11^ The Georgia SENDSS receives information on all cases with a notifiable disease including COVID-19 infection.

COVID-19 cases in the SENDSS database are subsequently grouped as confirmed or probable based on laboratory results. All the three data sources are housed at the Georgia Department of Public Health. Because the data were collected for public health surveillance purpose, this study has qualified for the Department’s Institutional Review Board exemption.

### Data Linkage

The three data sources were linked using a quasi-unique identifier – a 15-digit alphanumerical code called the Georgia Longitudinal ID (LONGID) which consists of letters from the first and last names, birthdate and a letter representing sex. The LONGID is collected by hospitals for the GCASR, and it was generated from available variables for the GRITS and SENDSS data; the three datasets were then linked with a deterministic linkage. Records without a valid LONGID value (0.03% of the GRITS records, 0.14% of SENDSS records, and 1.30% of GCASR records) were excluded from linkage.

### Outcome and Predictors

Acute ischemic and hemorrhagic stroke within 21 days post-initial vaccination were the outcomes of interest. The 21-day cutoff was chosen to ensure that patients were not yet eligible for a second dose of any of the available vaccines. Type of first vaccine dose was the main predictor and demographic variables such as age (grouped into 18–<45yrs, 45–<65yrs, 65– <80yrs, and 80+yrs), race (grouped as non-Hispanic white, non-Hispanic black, Hispanic and others), sex, and COVID-19 infection status were considered as covariates. A subject with a confirmed COVID-19 infection during the 21 days post-vaccination was considered as having a concurrent infection. Those who had confirmed infection prior to receiving the vaccine were grouped as having a past medical history of COVID-19 infection.

### Statistical Analysis

About 0.11% of the GRITS record had unspecified vaccine type and were excluded. Information on race was missing on 7.3% of the records as well. Missing values for the linking and predictor variables were assessed for randomness looking at the distribution of the records by other variables. Data were analyzed assuming the missingness was completely at random.

Logistic regression was applied to assess whether the relative risk, measured by odds ratio, of early stroke varies by vaccine type accounting for demographic variables and COVID-19 infection status. To determine if such risk would vary by the status of COVID-19 infection, we tested the statistical significance of an interaction term. All the analysis were done using SAS® Enterprise Guide 7.1 (Copyright © 2017, SAS Institute Inc., Cary, NC, USA)

## Results

About 5 million adult Georgians received at least one COVID-19 vaccine from December 1, 2020 to February 28, 2022. The median age was 51 years (IQR: 35-64), and the majority were white (51%) and female (55%) (see Table 1). Approximately 9% had COVID-19 infection prior to receiving the vaccine and 0.4% acquired the infection during the 21 days post-vaccination. 54% received the BNT162b2 vaccine, 41% received mRNA-1273, and 5% received Ad26.COV2.S.

**Table 1.** Description of Adult COVID-19 Vaccine Recipients and 21-day post vaccination incidence of Ischemic Stroke Among Adult Georgians, Dec 2020 – Feb 2022.

| **Characteristic** | **Total Vaccinated Adults**<br>**(N= 4,980,068)** | **Ischemic Stroke Within 21 Days**<br>**n (per 10<sup>5</sup>)** |
| --- | --- | --- |
| **Age (Year)** |  |  |
| 18-44 Yrs. | 1,973,482 (39.63) | 23 (1.165) |
| 45-64 Yrs. | 1,784,315 (35.83) | 134 (7.510) |
| 65-79 Yrs. | 977,600 (19.63) | 203 (20.765) |
| 80+ Yrs. | 24,4671 (4.91) | 113 (46.184) |
| **Gender** |  |  |
| Female | 2,739,302 (55.01) | 229 (8.360) |
| Male | 2,230,219 (44.78) | 244 (10.941) |
| Unknown | 10,547 (0.21) | -- |
| **Race** |  |  |
| Non-Hispanic Whites | 2,343,736 (50.75) | 217 (9.259) |
| Non-Hispanic Blacks | 1,245,790 (26.97) | 156 (12.522) |
| Hispanics | 336,711 (7.29) | 7 (2.079) |
| Non-Hispanic Others | 692,117 (14.99) | 57 (8.236) |
| **Previous<sup>†</sup> COVID-19 infection** |  |  |
| Yes | 446,252 (8.96) | 49 (10.980) |
| No | 4,533,816 (91.04) | 424 (9.352) |
| **Concurrent<sup>†</sup> COVID-19 infection** |  |  |
| Yes | 18,864 (0.38) | 14 (74.215) |
| No | 4,961,204 (99.62) | 459 (9.252) |

| **Vaccine Type of 1<sup>st</sup> Dose** |  |  |
| --- | --- | --- |
| Pfizer | 2,689,067 (54.00) | 219 (8.144) |
| Moderna | 2,037,360 (40.91) | 227 (11.142) |
| Johnson & Johnson | 248,184 (4.98) | 26 (10.476) |
| Unspecified | 5,457 (0.11) | 1 (18.325) |

N.B.:- $\phi$ : Individuals who were reported to the Georgia State Electronic Notifiable Disease Surveillance System (SENDSS) as confirmed COVID-19 infection prior to receiving the first COVID-19 Vaccine were considered as having previous infection Individuals who were reported to the Georgia SENDSS as confirmed COVID-19 infection during the 21 days post-first COVID-19 vaccine dose were considered as having concurrent infection

The 21-day post vaccination incidence of ischemic stroke was 8.14, 11.14, and 10.48 per 100,000 for BNT162b2, mRNA-1273 and Ad26.COV2.S recipients, respectively; after adjusting for age, race, gender, and COVID-19 infection status there was a 57% higher risk (OR=1.57, 95% CI: 1.02, 2.42) for ischemic stroke within 21 days of vaccination associated with Ad26.COV2.S vaccine compared to BNT162b2 (Table 2). There was no difference seen in risk of stroke between mRNA-1273 compared to BNT162b2. Those with concurrent COVID infection had an increased risk of ischemic (OR=8.00, 95% CI: 4.18, 15.31) and hemorrhagic stroke (OR=5.23, 95% CI: 1.11, 24.64). There was no statistical evidence for interaction between vaccine type and concurrent COVID infection.

**Table 2.** Relative Risk of Ischemic and Hemorrhagic Stroke Within 21 Days of Receiving the First Dose of COVID-19 Among Adult Georgians, Dec 2020-Feb 2022.

| Characteristic | Ischemic Stroke |  | Hemorrhagic Stroke |  |
| --- | --- | --- | --- | --- |
|  | Odds ratio (95% CL) | p-value | Odds ratio (95% CL) | p-value |
| **Age group** |  |  |  |  |
| 18 – <45 years | 0.03 (0.02, 0.04) | <.0001 | 0.12 (0.05, 0.31) | <.0001 |
| 45 – <65 years | 0.16 (0.12, 0.21) |  | 0.46 (0.22, 0.97) |  |
| 65 – <80 years | 0.45 (0.35, 0.58) |  | 0.88 (0.42, 1.85) |  |
| 80+ years | Reference |  | Reference |  |
| **Gender** |  |  |  |  |
| Female | 0.69 (0.57, 0.83) | 0.0001 | 0.86 (0.56, 1.33) | 0.51 |
| Male | Reference |  | Reference |  |
| **Race/Ethnicity group** |  |  |  |  |
| Non-Hispanic Black | 1.81 (1.47, 2.24) | <.0001 | 1.67 (1.05, 2.64) | 0.01 |
| Hispanic | 0.48 (0.23, 1.03) |  | 0.25 (0.04, 1.85) |  |
| Non-Hispanic Other | 1.12 (0.83, 1.50) |  | 0.54 (0.23, 1.28) |  |
| Non-Hispanic White | Reference |  | Reference |  |
| **COVID-19 Status** |  |  |  |  |
| Previous infection | 1.05 (0.73, 1.52) | 0.78 | 1.35 (0.65, 2.80) | 0.43 |
| Concurrent infection | 8.00 (4.18, 15.31) | <.0001 | 5.23 (1.11, 24.64) | 0.04 |
| **Vaccine type** |  |  |  |  |
| Johnson & Johnson | 1.57 (1.02, 2.42) | 0.13 | 1.09 (0.39, 3.05) | 0.73 |
| Moderna | 1.03 (0.85, 1.25) |  | 0.84 (0.54, 1.32) |  |
| Pfizer | Reference |  | Reference |  |

N.B.: § Hemorrhagic stroke includes intracerebral and subarachnoid hemorrhages

## Discussion

This statewide analysis showed that concurrent COVID-19 infection within the 21 days after vaccination was the strongest association with an increased risk of ischemic and hemorrhagic stroke regardless of vaccine type. An early increased risk of ischemic stroke within 21 days of vaccination with Ad26.COV2.S compared with BNT162b2 was also found after adjusting for age, race, gender, and COVID-19 infection status.

The early increased risk of both ischemic and hemorrhagic stroke in those vaccinated and with concurrent COVID-19 was independent of vaccine type. COVID-19 has been associated with an increased risk of acute ischemic stroke and intracerebral hemorrhage, and compared to stroke patients without infection, COVID-19 stroke patients tend to be younger and to have more severe strokes, a higher frequency of large artery occlusion and higher in-hospital mortality.^12^ This suggests that COVID-19 infection leads to a much higher risk of stroke outweighing the risk of stroke associated with vaccine alone.

COVID-19 vaccines using an adenoviral vector have been reported to have the rare occurrence of thrombotic thrombocytopenia including thrombosis of the cerebral veins.^3,13,14^ These cases have had positive heparin-PF4 HIT antibody ELISA tests in the absence of prior exposure to heparin. The FDA has reported that a causal relationship between Ad26.COV2.S vaccine and TTS is plausible, updating the EUA with a warning about rare clotting events after Ad26.COV2.S vaccination.^15^ In a self-controlled case series method study of early cardiovascular events after vaccine in France, the incidence of myocardial infarction was increased during the second week after vaccination with a single dose of Ad26.COV2.S however no association was seen with stroke.^10^ In France, the Ad26.COV2.S vaccine was limited to people aged 55 years or older whereas no age limit was present in Georgia which may have accounted for differences in increased rates of ischemic stroke seen compared to BNT162b2.^16^

A key strength of this study is the large registries that were able to link statewide vaccination date and type with statewide stroke diagnoses and COVID-19 test positivity. A limitation of this study is the lack of data on comorbidities that may have contributed to differences in stroke risk across vaccine recipients. Additionally, the increased use of home COVID-19 tests likely contributed to an underreporting of COVID-19 infection in the SENDSS database which may have influenced the study results. Finally, data was unavailable to determine if any of these early post-vaccination strokes were related to thrombotic thrombocytopenia.

Although not all determinants of stroke, particularly comorbidities, were considered in this analysis, concurrent COVID-19 infection had the strongest association with early incidence of ischemic and hemorrhagic strokes after first dose COVID-19 vaccine recipients compared with those without infection. The Ad26.COV2.S vaccine appears to be associated with higher risk of early post-vaccination ischemic stroke than BNT162b2 vaccine further supporting the ACIP preferential recommendation for mRNA COVID-19 vaccines over the Ad26.COV2.S vaccine.

## Data Availability

All data produced in the present work are contained in the manuscript

## Acknowledgments

**None**

